# Genomic Insights for Personalized Care: Motivating At-Risk Individuals Toward Evidence-Based Health Practices

**DOI:** 10.1101/2024.03.19.24304556

**Authors:** Tony Chen, Giang Pham, Louis Fox, Nina Adler, Xiaoyu Wang, Jingning Zhang, Jinyoung Byun, Younghun Han, Gretchen R.B. Saunders, Dajiang Liu, Michael J. Bray, Alex T. Ramsey, James McKay, Laura Bierut, Christopher I. Amos, Rayjean J. Hung, Xihong Lin, Haoyu Zhang, Li-Shiun Chen

## Abstract

**Background:** Lung cancer and tobacco use pose significant global health challenges, necessitating a comprehensive translational roadmap for improved prevention strategies. Polygenic risk scores (PRSs) are powerful tools for patient risk stratification but have not yet been widely used in primary care for lung cancer, particularly in diverse patient populations.

**Methods:** We propose the GREAT care paradigm, which employs PRSs to stratify disease risk and personalize interventions. We developed PRSs using large-scale multi-ancestry genome-wide association studies and standardized PRS distributions across all ancestries. We applied our PRSs to 796 individuals from the GISC Trial, 350,154 from UK Biobank (UKBB), and 210,826 from All of Us Research Program (AoU), totaling 561,776 individuals of diverse ancestry.

**Results:** Significant odds ratios (ORs) for lung cancer and difficulty quitting smoking were observed in both UKBB and AoU. For lung cancer, the ORs for individuals in the highest risk group (top 20% versus bottom 20%) were 1.85 (95% CI: 1.58 – 2.18) in UKBB and 2.39 (95% CI: 1.93 – 2.97) in AoU. For difficulty quitting smoking, the ORs (top 33% versus bottom 33%) were 1.36 (95% CI: 1.32 – 1.41) in UKBB and 1.32 (95% CI: 1.28 – 1.36) in AoU.

**Conclusion:** Our PRS-based intervention model leverages large-scale genetic data for robust risk assessment across populations. This model will be evaluated in two cluster-randomized clinical trials aimed at motivating health behavior changes in high-risk patients of diverse ancestry. This pioneering approach integrates genomic insights into primary care, promising improved outcomes in cancer prevention and tobacco treatment.

## Introduction

The worldwide burden of lung cancer and tobacco smoking presents major challenges to global health^1^. Evidence-based practices to reduce their risk such as cancer screening and tobacco treatment (e.g. smoking cessation medication) have long existed but are infrequently used in most primary care practices. Communication of the precision risk of lung cancer and precision benefit of smoking cessation is a promising but untested strategy to promote health behavior changes to reduce cancer risk. To address this gap, polygenic risk scores (PRSs) emerge as a valuable approach to assess disease susceptibility among populations and pinpoint individuals at higher risk^2–4^. PRSs can be derived from large-scale genome-wide association studies (GWAS) to estimate individual disease risk and have shown promise in predicting health outcomes and promoting preventive healthcare^5–9^. Despite their potential, PRSs’ implementation in primary care is limited, especially in diverse populations. Implementing a PRS-based precision intervention is crucial in order to address the multifaceted needs of different communities and individuals^10^. Harnessing PRSs effectively can make significant progress in mitigating lung cancer’s public health impact.

Ongoing studies like eMERGE^11^, GenoVA^12^, and WISDOM^13^ are leading the implementation of PRS into genetic risk reports (**Table 1**). They aim to personalize medical reports and understand the impact of PRS on screening, diagnostic procedures, and patient behavior. Notably, a gap persists as these initiatives have not yet formulated a PRS specifically for lung cancer. A likely reason is that the global burden of lung cancer is primarily driven by tobacco smoking rather than genetics^14,15^. However, accounting for the genetic basis of lung cancer may provide patients and clinicians with additional actionable information. The unique value proposition of a lung cancer-specific PRS lies in leveraging established and clear guideline-based prevention strategies, including smoking cessation treatment and lung cancer screening^16^. By incorporating PRSs for lung cancer and difficulty quitting smoking without treatment, there is an opportunity to revitalize and enhance these often-under-utilized prevention practices.

**Table 1.** Research on PRS use in clinical trials. We compare the PRECISE and MOTIVATE trials, part of our GREAT framework, with existing PRS-informed trials: GenoVA, eMERGE, and WISDOM. Bolded text in the PRECISE / MOTIVATE column highlight the points where our trials differ from the current trials. Namely, the PRECISE and MOTIVATE trials investigate lung cancer and smoking and will focus on <u>high risk</u> patients who are smokers or eligible for lung cancer screening. We also look at lung cancer screening, tobacco treatment, and smoking cessation as unique target outcomes. Finally, in addition to genetic and clinical risk messaging, the two trials have a unique emphasis on behavior mechanisms around lung cancer and smoking.

|  | GenoVA | eMERGE | Wisdom | PRECISE / MOTIVATE |
| --- | --- | --- | --- | --- |
| **Goal** | Motivate early diagnosis | Increasing clinical actions to mitigate risk of future disease | Personalized screening frequency | Motivate lung cancer screening (LCS) / tobacco treatment |
| **Targets** |  |  |  |  |
| **Target population** | All general Veterans' Affairs PCP* and patients | Patients in the large healthcare system | All women can sign up online | **High risk** PC patients (LCS eligible / smokers) and their PCPs |
| **Condition(s)\*\*** | BrCa, PrCa, CRCa, Afib, CAD, T2D | BrCa, PrCa, CRCa, Afib, CAD, T1D, T2D, BMI, Asthma, CKD, HCL | BrCa | **LC, smoking cessation** |
| **Target outcome** | New diagnoses | Change screening practice or lifestyle | Screening compliance and cancer detection | **LCS, tobacco tx, and smoking cessation** |
| **Polygenic Risk Scores** |  |  |  |  |
| **Genotyping** | Study team |  | Color genomics | 23andMe |
| **Imputation** | 1000 Genomes | All of Us |  | TOPMed |
| **Create PRS** | Top or many variants<br>European weights<br>Ancestry calibration | Terra cloud platform<br>Ancestry calibration | Color genomics | Top independent variants<br>Multi-ancestry weights<br>Ancestry calibration |
| **Risk Stratification** | 2 levels (OR of 2) | OR of 5-10 |  | 3 levels (top 20%, middle, bottom 20%) or tertiles |
| **Intervention Design** |  |  |  |  |
| **Risk representation** | Only genetic<br>Format of relative risk | Combined clinical + genetic<br>Format of absolute risk | Combined clinical + genetic<br>Format of absolute risk | Separate genetic + clinical<br>Format of relative risk |
| **Messaging** | Average vs. elevated risk | Average vs. elevated risk | No discussion of risk<br>Only recommend screening schedule | At risk, at high risk, or at very high risk |
| **Behavior mechanisms targeted** |  |  |  | **Perceived risk/benefit, Self efficacy, Personal relevance** |

\* PCP=primary care provider
\*\* BrCa=breast cancer, PrCa=prostate cancer, CRCa=colorectal cancer, Afib=atrial fibrillation, CAD=coronary artery/heart disease, T2D=type 2 diabetes, T1D=type 1 diabetes, BMI=body mass index/obesity, CKD=chronic kidney disease, HCL=hypercholesterolemia, lung cancer=lung cancer

We introduce the <u>G</u>enomic Informed Care for Motivating High <u>R</u>isk Individuals <u>E</u>ligible for Evidence-b<u>a</u>sed Prevention (GREAT) framework as a novel approach to incorporate PRS-enabled interventions in clinical settings (**Figure 1**). The core of GREAT is the use of PRSs that offer precise risk estimates for lung cancer and difficulty quitting smoking. By providing patients with personalized risk information, we aim to activate behavior change mechanisms that promote preventive actions. The primary targets of this intervention are high-risk individuals eligible for evidence-based prevention practices, such as lung cancer screening and smoking cessation. By integrating precision risk information with the benefits of timely interventions, GREAT empowers patients to make informed decisions about their health and motivates them to take proactive steps towards prevention. The personalized information may increase the patient uptake of evidence-based treatment by activating potential mechanisms such as treatment. Guided by enhanced outcome expectancy and self-efficacy based on the Capability-Opportunity-Motivation-Behavior (COM-B) model and Theoretical Domains Framework (TDF)^17–19^. Ultimately, our objective is not only to motivate, but to significantly reduce lung cancer morbidity and mortality through this innovative care paradigm.

**Figure 1.**
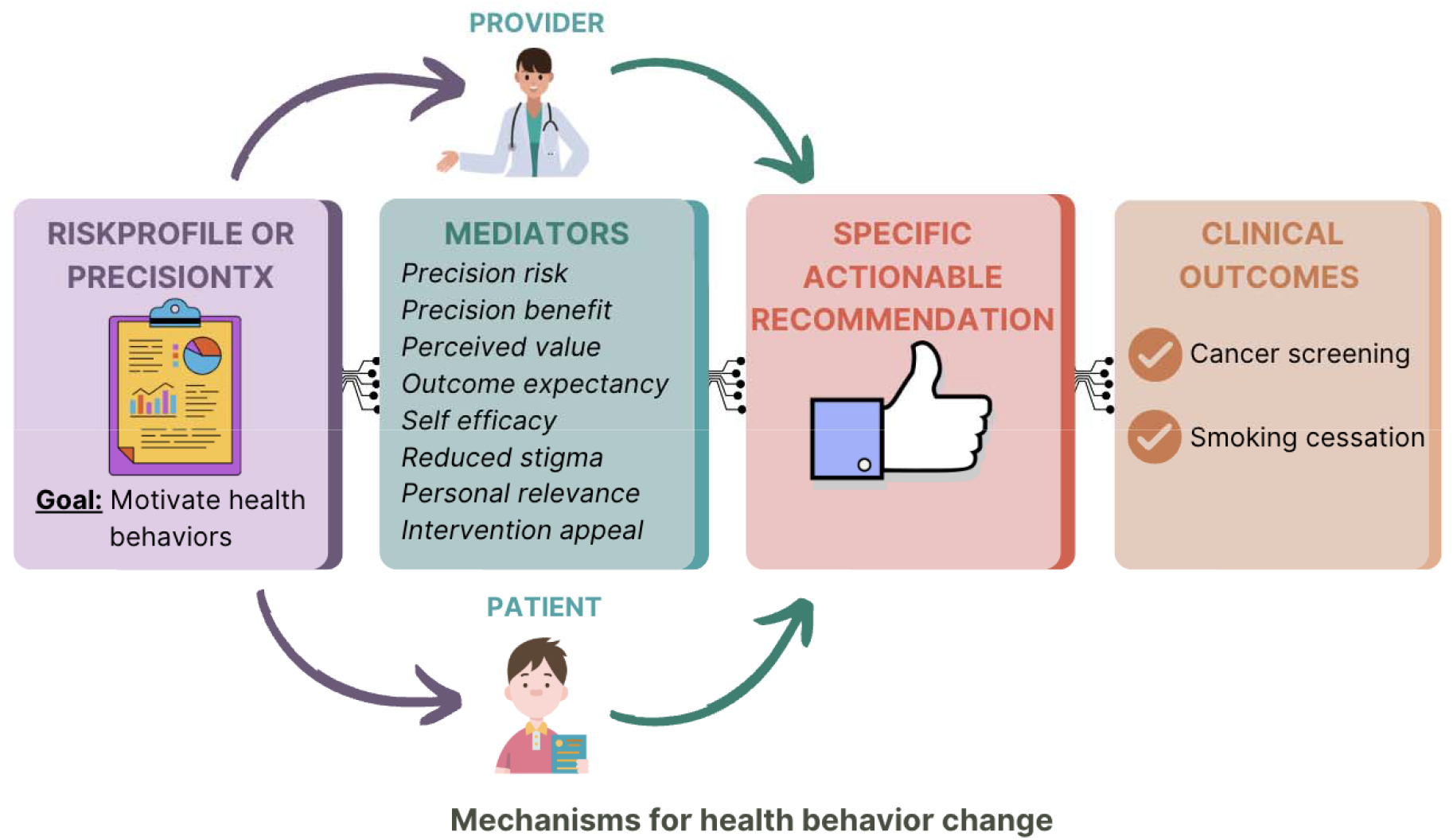
Care Paradigm: <u>G</u>enomic Informed Care for Motivating High <u>R</u>isk Individuals <u>E</u>ligible for Evidence-b<u>a</u>sed Prevention (GREAT). The GREAT framework is a primary care paradigm that integrates genetic and clinical risk in precision health. Individuals and their providers in two upcoming trials (PRECISE and MOTIVATE) are enrolled and provided with multilevel interventions (e.g. RiskProfile and PrecisionTx) to promote clinical outcomes of lung cancer screening, tobacco treatment, and successful smoking cessation in primary care settings. Mechanisms of health behavior changes (e.g., perceived benefit, self-efficacy, and outcome expectancy) will be evaluated. During the specific actionable recommendations phase, personalized shared decision-making will be facilitated by multilevel actions between patients and clinicians for better clinical outcomes.

Effectively translating PRSs into clinical practice requires a comprehensive and pragmatic translational roadmap for equitable and effective implementation (**Figure 1**). <u>First</u>, to address ancestry diversity, we take a two-step approach of (1) constructing PRS based on large-scale multi-ancestry GWAS, and (2) standardizing PRS distributions across the continuum of genetic ancestry to ensure accurate risk stratification based on PRS distributions for all patients^20^. <u>Second</u>, to document accuracy and transportability of the PRSs to diverse populations, we perform large-scale validation using diverse data from the Genetic Informed Smoking Cessation (GISC) trial^21^, UK Biobank (UKBB)^22^, and All of Us Research program (AoU)^23^. <u>Third</u>, we translate risk into actionable categories by setting appropriate thresholds for the PRS. The alignment of these thresholds with clinical significance involves many considerations for meaningful risk stratification. <u>Fourth</u>, we propose clear and patient-friendly communication strategies, including visual aids and educational materials, to facilitate understanding and meaningful interactions between patients and healthcare providers. Effectively communicating both the risk and precision of the PRS results is challenging but essential to empower patients to make informed decisions about their health. Moreover, it is crucial to consider patient perceived risk, perceived benefit, and personal relevance when discussing PRS results with patients. Patients’ understanding and interpretation of PRS may vary, leading to differing levels of engagement in preventive actions. Hence, comprehensive patient education programs can enhance awareness and knowledge about PRS, its implications, and available preventive measures.

In this paper, we introduce the design for two cluster randomized clinical trials (RCT): (1) PRECISE, which evaluates the effectiveness of a multilevel intervention, *RiskProfile*, on increasing lung cancer screening and tobacco treatment utilization in primary care (NIDA Grant 5R01CA268030-02); and (2) MOTIVATE, which evaluates the effect of PrecisionTx, a multilevel intervention to promote precision tobacco treatment in primary care (NIDA Grant 5R01DA056050-02). Through the innovative use of PRS, our aim is to motivate lung cancer screening and tobacco treatment among high-risk patients. We present a new care paradigm (**Figure 1**) and outline a translational roadmap (**Figure 2**) that discusses potential barriers and solutions for implementation. By incorporating personalized risk assessments, such PRS-enabled interventions have the potential to significantly improve lung cancer prevention strategies and patient outcomes.

**Figure 2.**
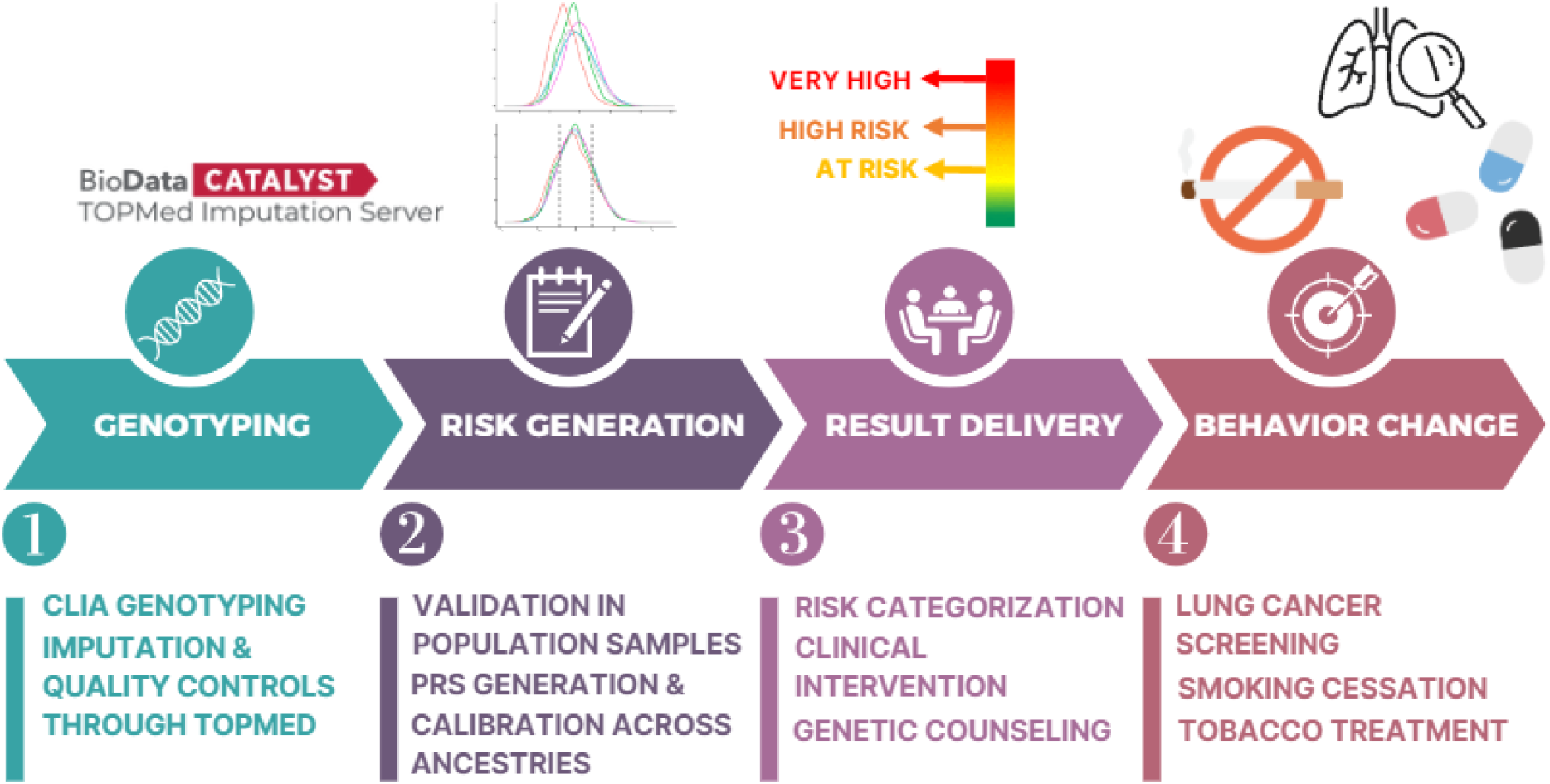
Roadmap for translating genetic data to a genetic risk profile as a multilevel intervention in primary care. In step 1, enrolled participants’ genetic data are analyzed by 23andMe’s Clinical Laboratory Improvement Amendments (CLIA) certified genotyping process. Imputation and quality controls are conducted through the Trans-Omics for Precision Medicine (TOPMed) server to ensure the integrity and reliability of the genetic data, as well as to impute the GWAS variants. Step 2 involves identifying available GWAS variants and weights to create the raw Polygenic Risk Scores (PRS). The PRS is adjusted for genetic ancestry using reference data such as the 1000 Genomes Project Phase 3 and applied to validation data such as the UK Biobank to establish risk categories and compute ORs. In step 3, these scores are converted into 3 risk levels based on the established thresholds. In step 4, a report with precision treatment is created and communicated to both the participant and the provider to make informed and educated decisions. Behavioral interventionists (research staff who are trained, certified, and supervised by a team of genetic counselor, psychologist, and psychiatrist) offer personalized guidance on behavior change, leveraging the updated genetic insights. The outcome aims to increase lung cancer screening orders, improve participant adherence, promote smoking cessation, and highlight the benefits of tobacco treatment.

## Materials and Methods

### 1000 Genomes Project Phase 3 reference data and principal components analysis

We use the 1000 Genomes Project Phase 3 (1000G)^24,25^ as a reference for genetic ancestry inference, as it is publicly accessible and includes genotype data from diverse populations. The 1000G dataset includes 3,202 individuals, with 633 Europeans (EUR), 893 Africans (AFR), 585 East Asians (EAS), 601 South Asians (SAS), and 490 Admixed Americans (AMR). We conducted principal components analysis (PCA) plink 2.0^26^ on all 3,202 samples, using 55,248 SNPs that are shared among the recommended SNPs set by gnomAD^27^, 1000G reference data, UKBB and GISC validation data, and the 23andMe genotyping array used for the trial (**Figure 3, Supplementary Figure 1, Supplementary Table 4**).

**Figure 3.**
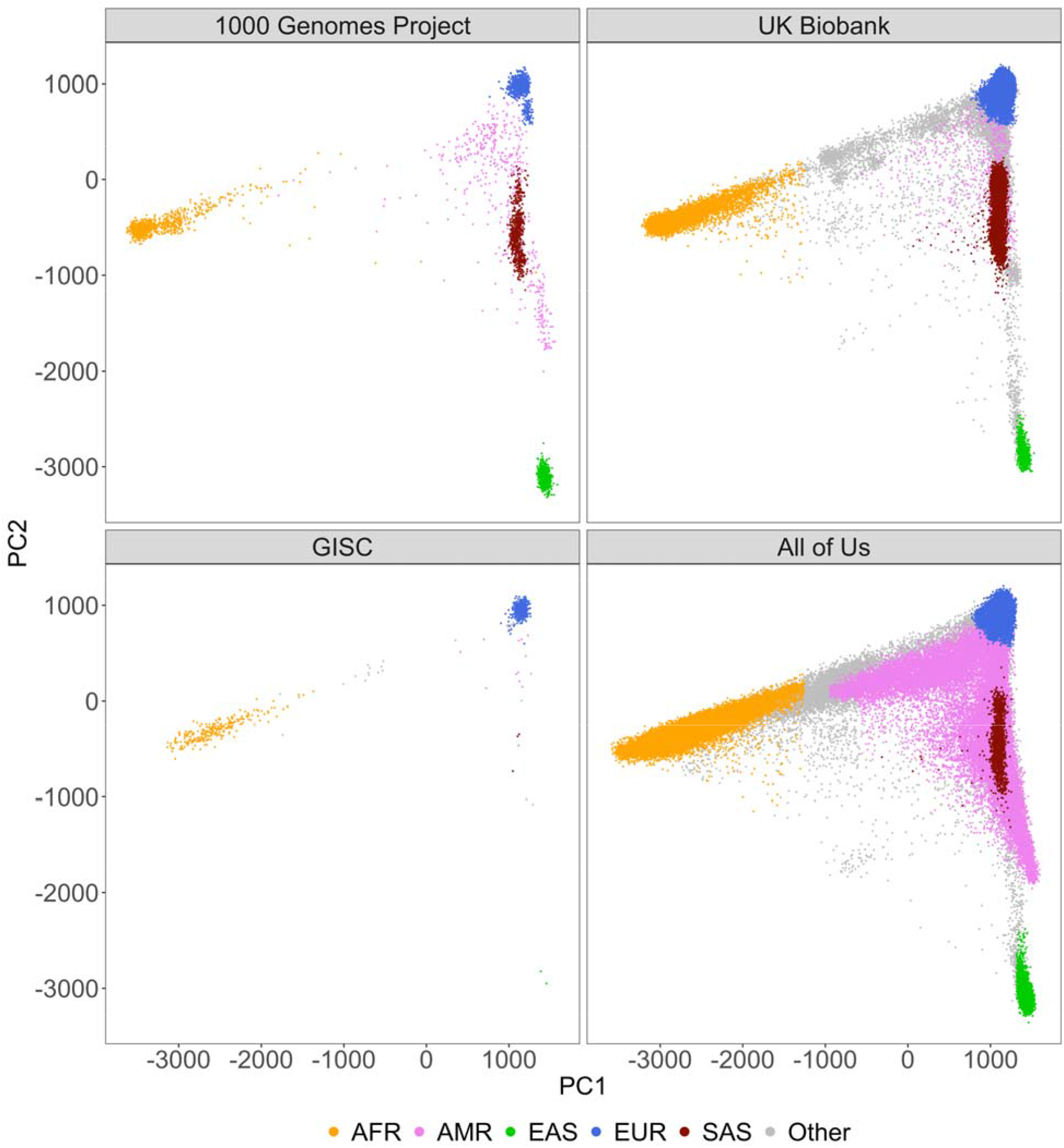
Cross-dataset distribution of genetic ancestry via PCA Projections in 1000G, GISC, UKBB, and AoU. This figure illustrates the utility of principal components analysis (PCA) loadings obtained from the 1000 Genomes Project Phase 3 (1000G) in discriminating ancestries within external datasets, the Genetically Informed Smoking Cessation (GISC) trial. PCA was initially conducted on the globally diverse genotype data of 1000G. The resultant PCA-space was then used to project genotype data from GISC, UKBB, and AoU. The scatter plot displays the first and second PCs for each individual in these datasets, with points distinctly marked by genetically-inferred ancestry.

We ran a random forest classifier to map each individual in 1000G to their respective populations using the first 5 PC scores. We then projected our validation data to this PC space and applied the same random forest classifier to produce “genetically-inferred” ancestry labels. Individuals with a predicted probability less than 90% for any of the five ancestries were labeled as “Other.” While our proposed framework removes the need for labels in the clinic, we use them here for illustrative purposes in validation.

### Validation in GISC, UK Biobank, and All of Us

We validated our PRS approaches in three datasets: Genetically Informed Smoking Cessation Trial (GISC)^21^, UK Biobank (UKBB)^22^, and All of Us Research Program (AoU)^23^ Here we present are brief background of the three studies. More details of the study background are provided in the **Supplementary Note**, and sample characteristics regarding sex, age, and outcomes information in UKBB and AoU are provided in **Supplementary Table 1.**

#### GISC Trial

The GISC trial, conducted at conducted at Washington University in St Louis^21^, included 822 current or previous smokers. Genetic data were available for 796 individuals (503 European, 257 African, 36 Other self-reported ethnicities), This dataset was primarily used for ancestry inference validation due to its relevance to our PRECISE and MOTIVATE trials.

#### UKBB

The UKBB dataset included 340,154 unrelated individuals with rich genetic and clinical data. This cohort consisted of 6,844 African, 730 Admixed American, 770 East Asian, 313,279 European, 7,197 South Asian, and 11,334 Other individuals by genetically-inference ancestry. The lung cancer analysis included 1,830 cases (ICD10 codes C34.0-C34.9) and 338,334 controls. The smoking cessation analysis involved 152,406 ever-smokers (117,483 former and 34,923 current), excluding 186,040 never smokers and 1,312 prefer-not-to-answer responses.

#### AoU

The AoU dataset included 210,826 unrelated individuals with whole-genome sequencing data. This cohort comprised 45,108 African, 32,563 Admixed American, 3,873 East Asian, 110,712 European, 1,689 South Asian, and 16,881 Other individuals by genetically-inference ancestry. We focused on 152,916 current or previous smokers for our smoking analysis.

### Construction of polygenic risk scores

Our trial incorporates the latest findings by utilizing recently genome-wide association study (GWAS) summary statistics for lung cancer^28^ and difficulty quitting smoking^29^, excluding UK Biobank samples to avoid overlapping with our validation data. While these meta-analyses predominantly consist of individuals of European ancestry, they also include a substantial proportion of non-European ancestry — about 26% for lung cancer and 21% for difficulty quitting smoking — which enhances the generalizability of the findings^30,31^.

For lung cancer risk, we started with 128 published SNPs found to be predictive of 5-year and lifetime cumulative risk for lung cancer^16^. Out of these, 101 SNPs overlapped with the published summary statistics, reference, and validation data (1000G, UKBB, GISC and AoU), and the 23andMe genotyping array used for the trial (**Supplementary Figure 1**). These SNPs were assigned effect sizes from the fixed-effect meta-analyses estimates in the latest lung cancer GWAS that includes EUR, AFR, and EAS ancestry^28^. For difficulty quitting smoking, we identified 175 SNPs predictive for smoking cessation following the same filtering procedure for lung cancer^29^.

The PRS construction began with the alignment of genotype data to the summary statistics, ensuring consistent PRS regardless of allele coding. Specifically, for any SNP *G* with reversed alleles, we recoded it as 2-*G* to avoid discrepancies. The raw PRS for an individual *i* with *M* SNPs was computed as

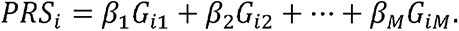

PRS calculations were performed using R, with genotype data input via the genio package^32^. PRS SNPs and weights for lung cancer and difficulty quitting smoking are provided in **Supplementary Tables 2-3**, respectively.

### Standardizing PRS distributions across the continuum of genetic ancestry

We standardized the PRS distributions for lung cancer and difficulty quitting smoking using the 1000G dataset, employing a regression-based method to adjust for distributional differences across ancestries (**Supplementary Tables 4-5**)^20^. This adjustment process involves two key steps:

#### 1. Mean adjustment

we conducted a linear regression of the raw PRS against the top five PCs derived from the PCA:

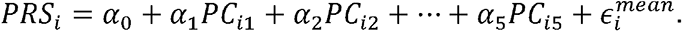

We then computed residuals *r_i_* of the raw PRS that account for mean differences in PRS distributions across ancestry:

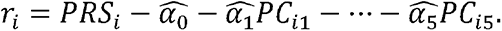

#### 2. Variance adjustment

using the square residuals 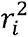 as a proxy for PRS variance, we ran a secondary linear regression:

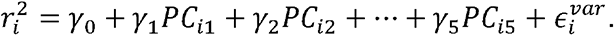

The final ancestry-adjusted PRS for each individual i was then computed as:

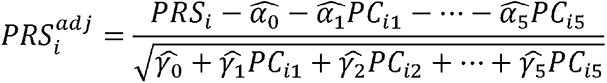

This standardization resulted in a PRS distribution with mean 0 and variance 1 across ancestries, ensuring that genetic risk is accurately reflected independent of ancestry. This method is crucial for individuals with admixed or unknown ancestry, where discrete ancestry-specific models are inappropriate^33,34^.

### Definition of patient risk categories

We stratified patients by genetic risk for lung cancer and difficulty quitting smoking by calculating odds ratios (ORs). ORs compare the probability of an outcome occurring in individuals within a percentile range *p* (i.e. 80-100%) of the ancestry-adjusted PRS distribution to those within a lower percentile range *q* (i.e. 0-20%).

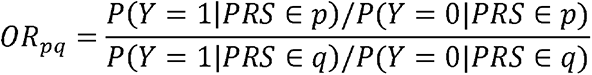

We established cut points in the PRS distribution to categorize individuals into three risk groups: “at risk”, “high risk”, and “very high risk”. For lung cancer, we defined these categories as the bottom 20%, middle 60%, and top 20% of the PRS distribution. For difficulty quitting smoking, categories were the bottom, middle, and top 33%. These thresholds were chosen to communicate personalized risks and benefits to high-risk patients in our trials, all of whom are eligible for lung cancer screening due to heavy smoking. Since difficulty quitting smoking lacks established absolute risk rates, we used these percentiles to provide a more agnostic risk assessment.

We use the distribution of our ancestry-adjusted PRSs among all 1000G samples to set percentile ranges and evaluate corresponding odds ratios among UKBB and AoU participants. For comparison, we also evaluate odds ratios using ancestry-matched raw PRS distributions, i.e. European-only 1000G PRS distribution for UKBB and AoU participants with genetically predicted European ancestry. For individual with “Other” predicted ancestry, we use the overall raw PRS distribution among all 1000G samples.

## Results

### Harmonization PRS distributions across ancestry

We found notable variation in raw PRS distributions across ancestries, highlighting that applying a universal cutoff for raw PRS without accounting for ancestry can lead to biased risk profiling and inaccurate clinical recommendations (**Figure 4**). However, our regression-based ancestry adjustment across all three datasets yields much more standardized distributions across ancestries. Specifically, the adjusted proportions of individuals within each risk category closely align with 20%-60%-20% for lung cancer, and 33.3%-33.3%-33.3% for difficulty quitting smoking. This adjustment ensures that patients of any background can be compared against a unified reference distribution for each outcome (**Supplementary Tables 6a-f**). This standardization places individuals across all ancestries on the same scale, allowing for a single risk stratification cutoff regardless of ancestral background. Such measures importantly enable fair risk assessment for individuals who may be labelled as “Other”, who may not fit well into a binned ancestry-specific risk model.

**Figure 4.**
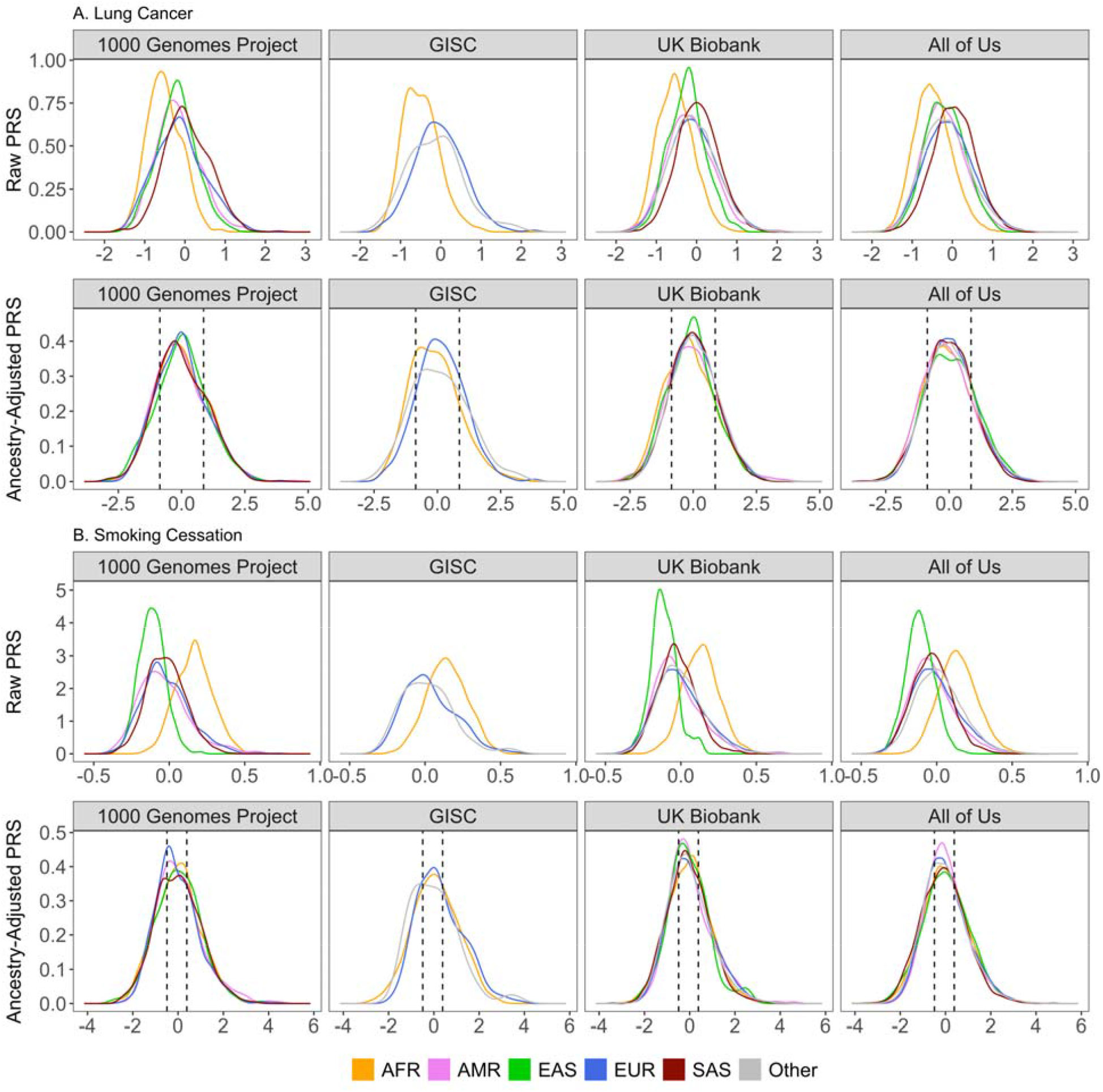
Ancestry adjustment of PRS for lung cancer and quit difficulty PRS across ancestral populations. We showcase the adjustment process for polygenic risk scores (PRS) for lung cancer (Panel A) and difficulty quitting smoking (Panel B) within the 1000 Genomes Project (1000G), GISC Trial, UK Biobank, and All of Us datasets. It displays both raw and ancestry-adjusted PRS, with data points color-coded according to genetically-inferred ancestries. EAS, AMR, and SAS ancestries were removed for GISC due to their small sample sizes. Ancestry adjustment effectively centers the PRS for different ancestries, mitigating the risk of incorrect stratification due to ancestry-related biases. Dotted vertical lines correspond to the 20^th^ and 80^th^ percentiles for lung cancer PRS distribution and 33^rd^ and 67^th^ percentiles for difficulty quitting smoking PRS among all 3,202 samples in the 1000 Genomes Project.

### Risk stratification for lung cancer and smoking cessation in UK Biobank and All of Us

After assigning UKB and AoU participants as “at risk”, “high risk”, or “very high risk” for lung cancer and difficulty quitting smoking, we identified significant odds ratios for both traits across different ancestry groups (**Figure 5**, **Supplementary Tables 7-10**). Our ancestry-adjusted PRS yielded similar ORs as using ancestry-matched distributions, demonstrating that a single PRS distribution can be appropriately applied to all individuals. In UKBB, the overall ORs for lung cancer 1.42 (95% CI: 1.24 – 1.65) for the “high risk” group and 1.85 (95% CI: 1.58 – 2.18) for “very high risk” group compared to the “at risk” group (**Supplementary Table 7a**). In AoU, these ORs were slightly higher – 1.65 (95% CI: 1.36 – 2.02) for “high risk” group and 2.39 (95% CI: 1.93 – 2.97) for “very high risk” group. While the ORs in non-European ancestries within UKBB were not significant, they were significant and even higher than those for European ancestries in AoU (**Supplementary Table 8a**). Moreover, the ORs for “Other” ancestry group in AoU were much higher and more significant using the ancestry-adjusted PRS compared to raw PRS, where an ancestry-matched risk distribution is least suitable (**Supplementary Table 8b)**.

**Figure 5.**
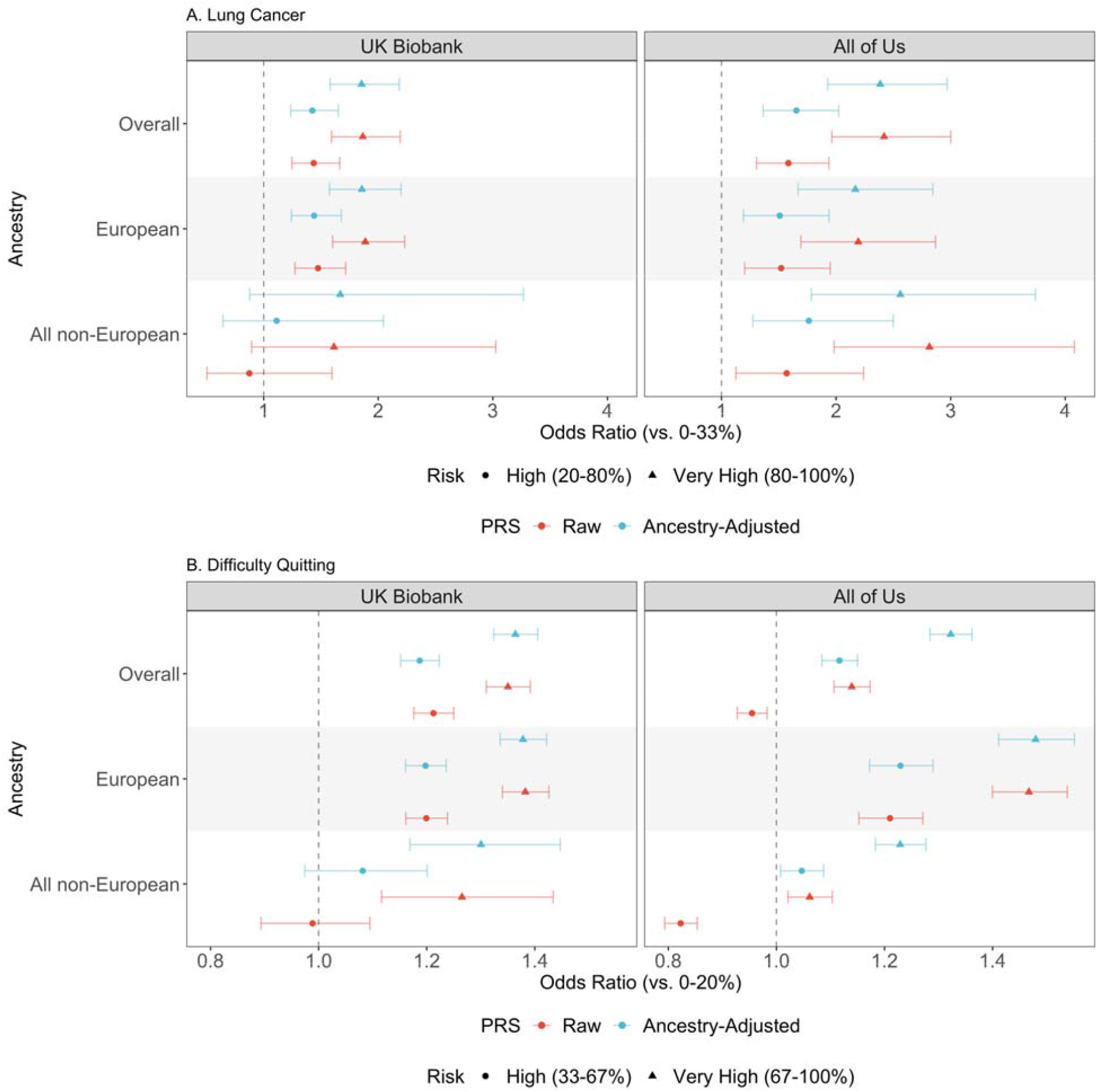
Risk stratification for lung cancer and difficulty quitting smoking using raw and ancestry-adjusted PRS. This figure illustrates the odds ratios (ORs) with associated 95% confidence intervals of PRSs for lung cancer (A) and difficulty quitting smoking (B) among UK Biobank (N=340,154 for lung cancer and N=152,406 for smoking), and All of Us (N=210,826 for lung cancer and N=152,916 for smoking) participants. We compare risk stratification using a raw PRS with ancestry-matched percentiles, and our ancestry-adjusted PRS with the same percentiles for all individuals.

We observed similar patterns for difficulty quitting smoking, where the ancestry-adjusted PRS performed similar or better than using ancestry-matched distributions for raw PRS. In UKBB, the overall ORs for difficulty quitting smoking were 1.19 (95% CI: 1.15 – 1.22) for “high risk” group and 1.36 (95% CI: 1.32 – 1.41) for “very high risk” group (**Supplementary Table 9a)**. In AoU, the ORs were 1.12 (95% CI: 1.08 – 1.15) for “high risk” group and 1.32 (95% CI: 1.28 – 1.36) for “very high risk” (**Supplementary Table 10a**) group. Notably, we observed higher and more significant ORs in non-European AoU participants when using the ancestry-adjusted PRS compared to the raw PRS. Although the improvement within each specific ancestry group was not as substantial, the aggregate ORs in all non-European groups increased from 0.89 (95% CI: 0.86 – 0.93) to 1.05 (95% CI: 1.01 – 1.09) in the “high risk” group, and from 1.07 (95% CI: 1.03 – 1.11) to 1.23 (95% CI: 1.18 – 1.28) in the “very high risk” group.

Using ancestry-adjusted PRS ensures accurate risk stratification across all ethnic backgrounds, a critical consideration given the substantial variability in raw PRS distributions across diverse populations. The outcome-based validation in UKBB and AoU further verifies that the ancestry-adjusted PRS provides valid risk stratification, and often yields better risk stratification for non-European ancestries compared to ancestry-matched modeling. These findings collectively facilitate a more robust and standardized application of PRS in clinical reporting.

### Translating polygenic risk scores into clinical reports

We highlight two example trials: PRECISE (NIDA Grant 5R01CA268030-02) and MOTIVATE (NIDA Grant 5R01DA056050-02), designed to promote health behavior change using genetically-informed interventions, RiskProfile and PrecisionTx, respectively. These interventions incorporate PRS to communicate precision risk of lung cancer and precision benefits of smoking cessation, promoting evidence-based practices such as cancer screening and tobacco treatment in high-risk individuals who smoke and/or are eligible for lung cancer screening. PRS risk stratification from either RiskProfile or PrecisionTx, along with clinical information, is delivered within a comprehensive report that includes actionable recommendations to reduce lifetime risk (**Figure 6** and **Figure 7**). Access to 23andMe genotypes and expanded health information has been a motivating component for the research participants. Both PRECISE and MOTIVATE are currently in the preliminary phases of recruiting primary care providers and patients. The recruitment strategy aims to engage over 100 physicians and 1600 patients in these trials.

**Figure 6.**
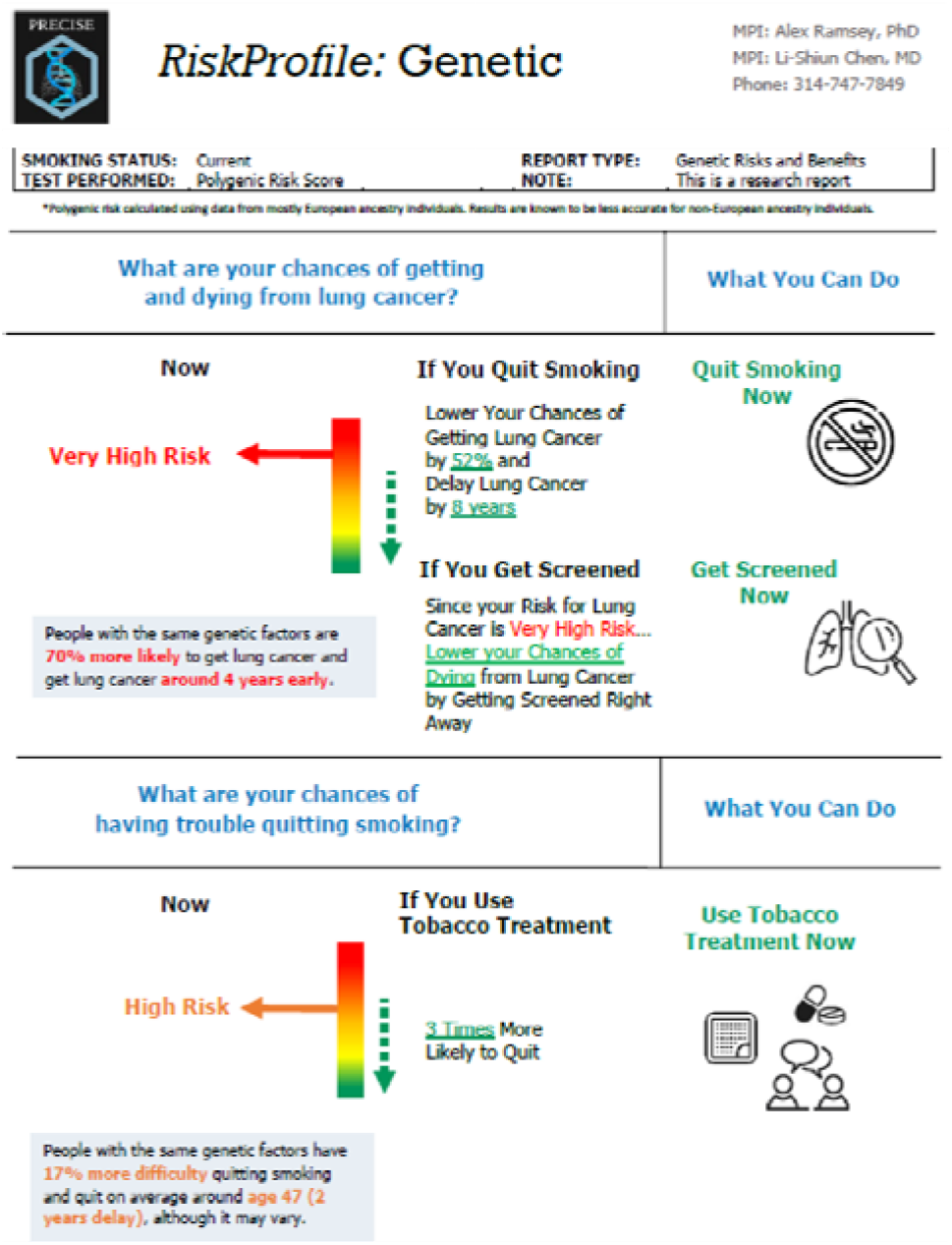
Example 1 clinical report: RiskProfile. We present example 1 for genomically-informed interventions using the GREAT framework. RiskProfile is designed to motivate lung cancer screening and tobacco treatment among screening-eligible patients. This intervention utilizes ancestry-adjusted PRS to stratify patients into “at risk” (yellow), “high risk” (orange), and “very high risk” (red) genetic risk categories. RiskProfile focuses on prevention and expands beyond personalized risk to also provide personalized benefit of cancer screening and use a multilevel intervention design directed to both physicians and patients in clinical settings. In an ongoing NCI-funded trial (NCT05627674), the effect of RiskProfile on clinician ordering and patient completion of lung cancer screening will be evaluated.

**Figure 7.**
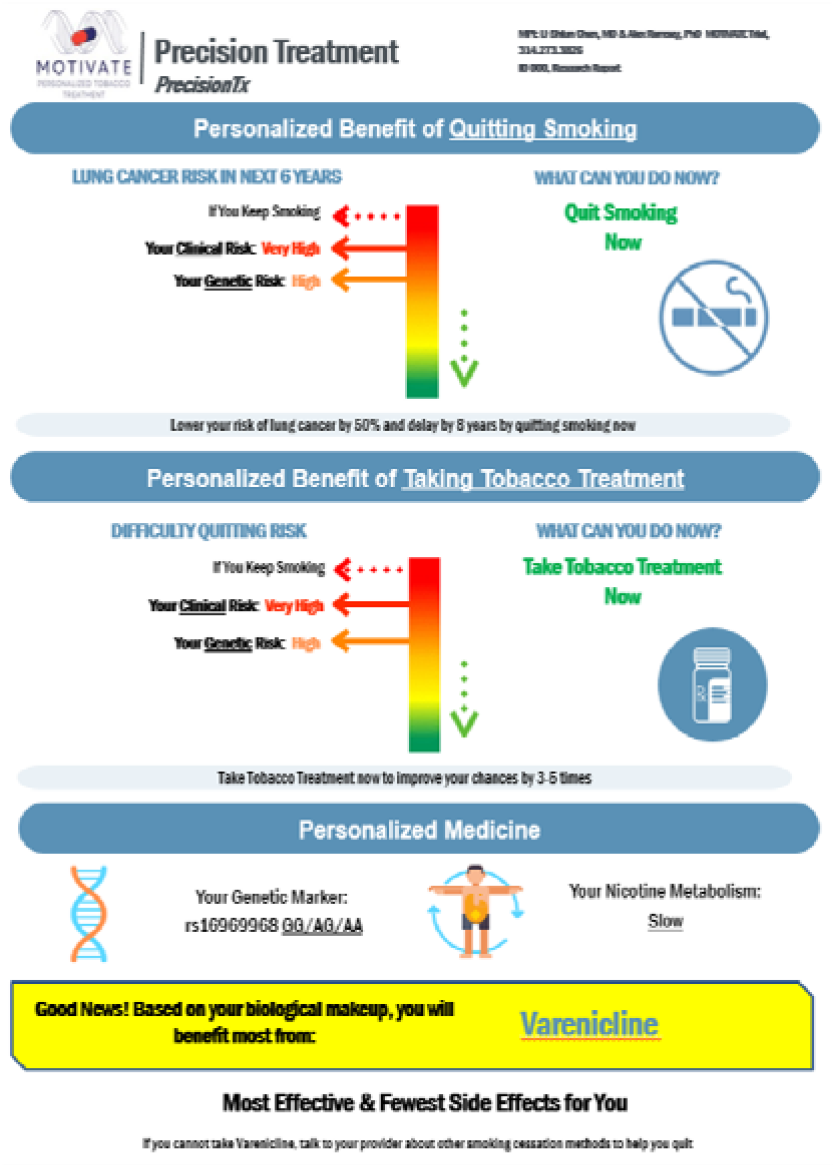
Example 2 clinical report: PrecisionTx. We present example 2 for genomically-informed interventions using the GREAT framework. PrecisionTx is designed to motivate tobacco treatment among patients who smoke. This intervention utilizes ancestry-adjusted PRS to stratify patients into “at risk” (yellow), “high risk” (orange), and “very high risk” (red) genetic risk categories. PrecisionTx focuses on treatment and expands beyond personalized risk to also provide personalized benefit of tobacco treatment, and use a multilevel intervention design directed to both physicians and patients in clinical settings. In an ongoing NIDA-funded trial (NCT05846841), the effect of PrecisionTx on clinician ordering, patient adherence, and smoking abstinence will be evaluated.

## Discussion

In this study, we introduce the GREAT framework in primary care, applying PRSs in two trials to provide precise risk estimates for lung cancer and difficulty quitting smoking to high-risk individuals. These estimates aim to activate behavior change mechanisms that promote health. To enable the interventions, we present a translational roadmap to implement genetic data in PRS-enabled interventions designed to promote health behaviors, evaluating their effectiveness in motivating high-risk patients to increase cancer screening and smoking cessation.

Importantly, we have accomplished our goals of generating behavior-change interventions by a) framing our translational message specifically for high-risk patients who have not received guideline-recommended cancer screening or tobacco treatment^35–38^, b) translating risk categories into precision risk and benefit that are designed to motivate health behavior changes, and c) ensuring inclusion of diverse ancestry with PC-regression-based PRS adjustment.

Our goal is to enable a robust implementation of PRS in currently funded clinical trials to evaluate the efficacy of these relatively novel interventions. The GREAT framework guides the implementation of PRS-enabled interventions in primary care. Future research need to address critical questions such as timing, methodology, and location of these interventions’ delivery to patients and providers to optimize its acceptability, understanding, and impact.

Our approach to ancestry adjustment of PRS employs widely accessible data from the 1000G dataset, as an alternative to methods in the GenoVA^12^ and eMERGE^11^ studies that use data from the Mass-General Brigham Biobank and AoU, respectively. We validated the transferability of our 1000G-based standardization in external datasets from the UKBB, GISC and AoU, allowing future trials to adopt a similar methodology irrespective of their specific genetic data. By utilizing our provided PC loadings and PRS standardization formula for lung cancer and difficulty quitting smoking, new patients in these trials can receive accurate risk categorization reports, bypassing the inaccuracies of self-reported ethnicity and the need for re-training PCA models.

A notable gap in current practice is the absence of genetic information in electronic health records (EHRs) for decision support and the lack of PRS generation in clinical labs. Implementing precision interventions in primary care necessitates a workflow that incorporates EHRs for recruitment, biomarker testing protocols, and standardized processes to generate personalized intervention reports^39^. This requires collaborations with primary care stakeholders, community advisory boards, genetic counseling, and health communication to improve intervention clarity, accuracy, and impact^40–44^. Patients expressed a notable interest in receiving personalized interventions. In our previous study, 85% of smoking patients reported a high interest in receiving genetically tailored tobacco treatment^45^. Further, a substantial majority (95%) of individuals who smoke endorsed the importance of receiving genetic results to guide their treatment^41^. Following receipt of a personalized genetic risk profile for smoking cessation, 91% of participants who smoke found the tool highly useful for understanding their health, cope with health risks, and feeling more in control of their health^42^. Such pronounced interest and the perceived significance of genetic data highlight the growing demand for personalized interventions among patients who smoke. Personalized interventions may increase patient compliance. For example, study found that patients reported higher interest in taking medication (97.5% vs. 61.0%, p<.0001) when medication was personalized based on their genetics^45^. However, substantial evidence suggests the complex interplay of fear, fatalism, and risk perception when patients face lung cancer screening^46–49^ Therefore, it is important to evaluate the translation potential of personalized genetics in motivating patients for positive behavior change.

Unlike most current research that evaluates PRS-enabled interventions in general patient populations, our work uniquely focuses on designing and evaluating these interventions specifically among high-risk patients who will benefit tremendously from recommended health behaviors such as lung cancer screening and smoking cessation, when general medical advice is not enough to motivate such behaviors.

Here we share three key design considerations for best practices. First, f<u>or equitable</u> <u>implementation</u> of precision health interventions, tools must be designed with active engagement from racial/ethnic minority communities, rather than solely examining their effectiveness for these communities post hoc. We engage in ongoing participatory sessions with racial and ethnic minority communities and advisory boards across all phases through iterative cycles of intervention development, feedback, and testing so that innovative genomics-informed tools are beneficial across diverse populations.

Second, we have chosen <u>clinically meaningful thresholding</u> for PRS risk categories in communicating personalized risks and benefits to patients. These categories were selected because all participants in our ongoing trials are high-risk primary care patients eligible for lung cancer screening, who are current or former heavy smokers. We defined lung cancer risk by the bottom 20%, middle 60% and top 20%, and difficulty quitting smoking by slightly more agnostic tertiles, to motivate positive behavior change. We aim to follow best practices of communicating uncertainties and deciding between a continuous or categorical risk presentation. Transparency about the imprecision in both risk estimates and action thresholds for PRS is essential.

Third, we plan to <u>update our workflow</u> to adapt to new GWAS findings and evolving methodologies. As scientific knowledge advances, outdated PRS predictions can hinder effective implementation. A dynamic PRS framework that allows for regular updates based on new discoveries is essential. This ensures that PRS-based interventions remain current and aligned with the latest advancements, reducing the implementation gap and maximizing their impact on preventive healthcare outcomes. Following current genetic counseling recommendations, we have established a process to incorporate new evidence into our intervention for smoking cessation and lung cancer risk. This process will adjudicating new population-specific evidence on genetics and biomarkers, evaluating its impact on personal and population-level risk changes, and effectively communicating the dynamic nature of genetic evidence to patients and providers.

Our work have several limitations, but we hope contribute to the knowledge pool for the best practices in creating PRS-enabled interventions that may be disease-, population-, or context-specific. Our approach is tailored for our unique outcomes (lung cancer and difficulty quitting smoking), population (patients eligible for lung cancer screening and tobacco treatment), and context (primary care) to optimize health impact. A notable limitation is the underrepresentation of non-European populations in the multi-ancestry GWAS employed to derive the PRS weights. This underrepresentation may reduce the predictive power of the PRS in non-European populations. The dominance of European populations persists in existing GWAS^50–53^, not limited to lung cancer and smoking cessation. However, with the burgeoning emphasis on incorporating minority populations in GWAS analyses and the development of new PRS approaches and toolkits to enhance predictive power in diverse populations ^7–9,34,54–56^, we can iteratively refine PRS implementation in our trial to synchronize with the latest advancements.

Many questions remain for the near future. First, <u>how can we reduce the time lag from</u> <u>evidence to implementation?</u> The constant evolution of evidence identifying new biomarkers for treatment is a challenge. For instance, our recent work highlights the potential of PRSs in guiding future treatment approaches^57^. Despite actionable precision treatment findings, the ever-changing evidence base and the perception of better data have hindered effective implementation^43,58^. To address this, we aim to use cutting-edge metabolic and genetic markers that offer robust evidence for precision treatment, reducing the time from evidence generation to practical application. This approach allows us to measure and report the time from landmark publications to the implementation of key findings ^59^, enhancing the efficiency and effectiveness of precision treatment and ensuring patients benefit from the latest recommendations.

<u>Second, can we truly evaluate the effect of precision interventions?</u> We expect that precision interventions may activate multiple mechanistic pathways to the uptake and efficacy of lung cancer screening or tobacco treatment. Understanding these mechanisms is essential to refine for intended outcomes and contexts. <u>Third, can these</u> <u>precision interventions be scaled in the real-world clinics?</u> Evidence shows that physicians are highly receptive to medication recommendations based on biomarkers^60^. To reduce burden, we need to leverage existing EHR tools (e.g. Best Practice Advisories) and training to efficiently facilitate physician prescribing^61,62^. Understanding of mechanistic and implementation outcomes will guide scalable, efficient delivery components for integration into clinic workflows^39^, using trained embedded staff, and digital therapeutic tools to enable these PRS-informed behavioral interventions^63^.

In conclusion, a well-designed roadmap that validates the PRS, incorporates multi-ancestry GWAS-based weights, translates risk into actionable categories, communicates effectively, considers patient perspectives, and accommodates evolving science is essential for the equitable and pragmatic translation of PRS into clinical care. By addressing the barriers and implementing potential solutions at each stage, we can leverage PRS to improve preventive healthcare and significantly reduce the burden of lung cancer.

## Data Availability

Relevant data is available online at https://github.com/chen-tony/GREAT.

## Acknowledgements

We would like to thank Reeya Joseph for her editorial support with the introduction, Peter Kraft for his advice on our manuscript, and Scott Vrieze for his assistance with summary statistics for difficulty quitting smoking. We would also like to thank and acknowledge the participants enrolled in the UK Biobank (obtained under UK Biobank resource application 52008) and GISC trial for contributing vital data to this work.

This research was supported by NIH Training Grant T32GM135117 and NSF Graduate Research Fellowship DGE-2140743 (T.C.), R35-3CA197449, R01-HL163560, U01-HG009088, and U01-HG012064 (X.L.), NIH Intramural Research Program (H.Z.), NIH 5T32-HL007776-25, R01-DA056050, R01-CA268030, P30-CA091842-19S5, P30-CA091842-16S2 and P50-CA244431 (L.C.) and U19-CA203654.

## Author Contributions

T.C., H.Z., and L.C. conceived the project, T.C., G.P., and L.F. carried out all data analyses under the supervision of H.Z. and L.C. G.S. and D.J. assisted with obtaining and processing GWAS summary statistics for difficulty quitting smoking. T.C., G.P., H.Z., and L.C. drafted the manuscript. All co-authors reviewed and approved the final version of the manuscript.

## Data and Code Availability

R code and plink commands, as well as accompanying data, used for analysis are provided in a walkthrough available on GitHub at https://github.com/chen-tony/GREAT.

Genotype data from the 1000 Genomes Project (Phase 3) were acquired via the plink website https://www.cog-genomics.org/plink/2.0/resources. Population information were obtained from public resources: ftp://ftp.1000genomes.ebi.ac.uk/vol1/ftp/release/20130502/ and https://ftp.1000genomes.ebi.ac.uk/vol1/ftp/data_collections/1000G_2504_high_coverage/1000G_698_related_high_coverage.sequence.index.

