## Supplementary Figure for "Genomic Insights for Personalized Care: Motivating At-Risk Individuals Toward Evidence-Based Health Practices"

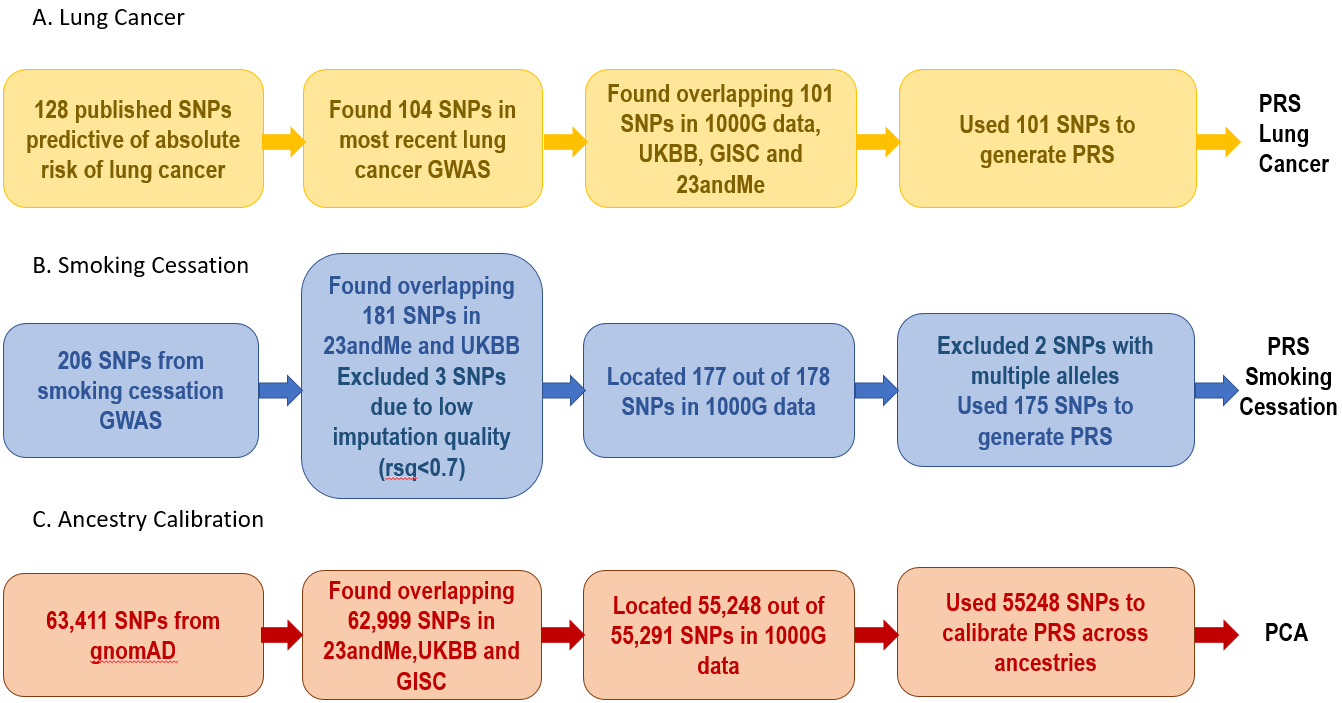

Supplementary Figure 1. Selection Process of Variants used in the PRS generation and Ancestry Adjustment. Variants identified in PRSs from large multi-ancestry GWAS for lung cancer^15,47^ (A) and smoking cessation^48^ (B). Variants identified in the principal components analysis (PCA) to standardize PRS distributions across ancestries (C). To construct PRS, we start with published sets of predictive SNPs, subsetted to those with available GWAS summary statistics and overlapping with the 23andme genotyping array and our ancestry adjustment and validation data. For PCA, we use a subset of established ancestry-informative SNPs^49^ that overlapped with the genotyping array and validation data.
