## Supplementary Note for "Genomic Insights for Personalized Care: Motivating At-Risk Individuals Toward Evidence-Based Health Practices"

**Study information about GISC, UK Biobank and All of Us**

**GISC Trial**: The Genetically Informed Smoking Cessation (GISC) trial is a prospective, randomized, placebo-controlled trial conducted at Washington University in St. Louis. This study includes 822 current or previous smokers. We focused on 796 individuals with genetic data: 647 current and 149 former smokers. The cohort comprised 503 individuals of European self-reported ethnicity, 257 of African self-reported ethnicity, and 36 self-reported as "Other". GISC data was utilized for ancestry inference validation, as this patient population closely resembles those expected in our PRECISE and MOTIVATE trials.

**UK Biobank (UKBB)**: The UKBB, a widely recognized dataset, provides extensive genetic and clinical data from approximately 500,000 British individuals. Our study included 340,154 unrelated individuals, up to third-degree relatives. The cohort consisted of 6,844 African, 730 Admixed American, 770 East Asian, 313,279 European, 7,197 South Asian, and 11,334 Other ancestry individuals. Participants had a mean age of 56.6 years (SD 8.2) and 54.1% were female (183,969 individuals). Our lung cancer analysis included 1,830 cases defined by at least one ICD10 code C34.0-C34.9 under Field 40006, and 338,334 controls with no recorded ICD10 codes. The smoking cessation analysis involved 152,406 ever-smokers, comprising 117,483 former and 34,923 current smokers, defined by difficulty quitting based on Field 20116. We excluded 186,040 never smokers and 1,312 participants who preferred not to answer.

**All of Us Research Program (AoU)**: The AoU is a biobank dataset dedicated to including individuals from diverse and historically under-represented backgrounds, with comprehensive health records for hundreds of thousands of individuals. We focused on 210,826 unrelated individuals with whole-genome sequencing data. The cohort included 45,108 individuals with African inferred ancestry, 32,563 Admixed American, 3,873 East Asian, 110,712 European, 1,689 South Asian, and 16,881 Other. Our smoking analysis was restricted to 152,916 individuals reported as current or previous smokers.
